# Long-Term Comparison of Cilostazol versus Clopidogrel in Secondary Prevention of Noncardioembolic Ischemic Stroke

**DOI:** 10.1101/2023.02.05.23285505

**Authors:** Yu Jeong Lee, Nam Kyung Je

**Author notes:** Corresponding author: Nam Kyung Je, College of Pharmacy, Pusan National University, Busandaehakro 63 Bungil 2, Geumjeong-Gu, Busan, 46241, Republic of Korea.

## Abstract

**Background:** Cilostazol is a widely used antiplatelet drug for secondary stroke prevention in Asia, but its comparison with clopidogrel, another commonly used antiplatelet drug, is not well understood. This study aims to investigate the effectiveness and safety of cilostazol compared to clopidogrel for the secondary prevention of noncardioembolic ischemic stroke.

**Methods:** A retrospective comparative effectiveness research analysis was conducted using 1:1 propensity-matched data from insured individuals between Jan 1, 2012, and Dec 31, 2019. The study used administrative claims data in Health Insurance and Review Assessment in Korea. Patients with diagnosis codes for ischemic stroke without atrial fibrillation, heart failure, valvular diseases, and myocardial infarction were included and divided into two groups, those receiving cilostazol and those receiving clopidogrel. The primary outcome was a recurrent ischemic stroke. Secondary outcomes included all-cause death, myocardial infarction, hemorrhagic stroke, and a composite of these outcomes. The safety outcome was major gastrointestinal bleeding.

**Results:** A total of 4754 patients were included in the propensity-matched population, with a median age of 67 years, 3080 (40.7%) were male, and 4480 (59.3%) were female. The study found no statistically significant difference in recurrent ischemic stroke (cilostazol group vs clopidogrel group, 2.7% vs 3.2%; 95% CI, 0.62-1.21), the composite outcome of recurrent ischemic stroke, all-cause death, myocardial infarction, and hemorrhagic stroke (5.1% vs 5.5%; 95% CI 0.75-1.22), and major gastrointestinal bleeding (1.3% vs 1.5%; 95% CI 0.57-1.47) between cilostazol and clopidogrel use patients. In subgroup analysis, cilostazol was associated with a lower incidence of recurrent ischemic stroke compared to clopidogrel in hypertensive patients (2.5% vs 3.9%; interaction P = 0.41).

**Conclusions:** This real-world, comparative effectiveness research study suggests that cilostazol is effective and safe for noncardioembolic ischemic stroke and may be associated with better effectiveness in hypertensive patients compared to clopidogrel over an 8-year period.

## Introduction

Patients who have had a noncardioembolic stroke or transient ischemic attack (TIA) are at increased risk for recurrent events.^1^ To reduce this risk, secondary prevention strategies, such as antiplatelet therapy, are crucial. Various antiplatelet agents, including clopidogrel, aspirin, aspirin-dipyridamole, cilostazol, and ticagrelor, can be applied to prevent secondary stroke events. For individuals with high-risk TIA, minor ischemic stroke, or stroke caused by intracranial large artery atherosclerosis, it is recommended to undergo a short-term dual antiplatelet therapy (DAPT) regimen of aspirin and clopidogrel for up to 90 days. Beyond 90 days post-stroke, single antiplatelet therapy (SAPT) is advised, as prolonged use of DAPT is associated with an increased risk of bleeding.^2–4^

Clopidogrel is one of the first-line antiplatelet agents for secondary prevention in patients with noncardioembolic ischemic stroke. It has been shown to be more effective than aspirin in preventing ischemic stroke, myocardial infarction, or vascular death.^5,6^ Additionally, the risk of gastrointestinal bleeding with clopidogrel is lower than that of aspirin.^6,7^

Cilostazol, an inhibitor of phosphodiesterase (PDE) III, increases cyclic adenosine monophosphate leading to reversible inhibition of platelet aggregation, vasodilation, and inhibition of vascular smooth muscle cell proliferation.^8^ It has been approved and widely used for secondary stroke prevention in Asian countries.^9^ However, in the United States and Europe, its use for secondary prevention of ischemic stroke is off-label and less common.^9^

Cilostazol has been found to be non-inferior to aspirin in preventing ischemic stroke through several randomized controlled trials in Asians.^10–12^ The Cilostazol versus Aspirin for Secondary Ischaemic Stroke Prevention (CASISP) trial in China showed that the hazard ratio (HR) of any stroke recurrence was 0.62 (95% confidence interval (CI), 0.30-1.26; P = 0.185) in cilostazol group compared to aspirin group and there were fewer brain bleeding events in the cilostazol group.^10^ Similarly, in the Cilostazol for Prevention of Secondary Stroke (CSPS) 2 study conducted in Japan for 2,716 patients, the cilostazol group was non-inferior to the aspirin group in terms of stroke recurrence (HR, 0.743; 95% CI, 0.564-0.98; P = 0.0357) and had fewer hemorrhagic events (HR, 0.458; 95% CI, 0.296-0.711; P = 0.0004).^11^ However, it is worth noting that the Prevention of Cardiovascular Events in Ischemic Stroke Patients with High Risk of Cerebral Hemorrhage (PICASSO) study did not find that cilostazol reduced the risk of hemorrhagic stroke (HR, 0.65; 97.5% CI, 0.27-1.57; P = 0 55).^12^

While some clinical trials have compared cilostazol with other antiplatelet agents, few have directly compared cilostazol with clopidogrel for secondary stroke prevention. According to the meta-analyses of randomized trials comparing cilostazol with placebo or other antiplatelet therapies, cilostazol monotherapy was superior to aspirin monotherapy in terms of efficacy and safety, but it is difficult to conclude its comparison with clopidogrel.^13–15^

We aimed to compare the long-term effectiveness and safety between cilostazol and clopidogrel as antiplatelet therapy for secondary prevention in patients with noncardioembolic ischemic stroke in a real-world setting.

## Methods

### Data source

We performed a retrospective analysis of the Korean Health Insurance and Review Assessment (HIRA) database, which comprises health insurance claims from the entire population of South Korea. The database contains information on patients’ demographics; diagnostic codes based on the International Classification of Diseases, 10th Revision; and details of medical examinations and treatments, such as diagnostic tests, procedures, surgeries, and drug prescriptions.^16^ The data used for this study were obtained from the HIRA research database (M20220628003) and were stored on a separate server managed by the HIRA. All authors had full access to all the data in the study and take responsibility for its integrity and the data analysis. This study protocol was approved by the institutional review board of the Pusan national university (No. PNU IRB/2022_114_HR) with a waiver of informed consent.

### Study population

We utilized the HIRA claims database to identify the initial hospitalization episodes for acute ischemic stroke for each patient between January 1, 2012, and December 31, 2019, ensuring that no prior diagnosis of ischemic stroke had been recorded within the preceding year. To qualify as an episode of acute ischemic stroke, the main diagnosis must be an ischemic stroke, and the patient must have undergone brain computed tomography (CT), brain magnetic resonance imaging (MRI), endovascular treatment (EVT), or intravenous thrombolytic therapy (IVT) during their hospital stay. This definition referenced an algorithm that identified acute ischemic stroke by comparing claims data and the stroke registry database in Korea.^17^

We established the following exclusion criteria: patients who expired during hospitalization, were transferred to another hospital upon discharge, were discharged the same day of admission, were hospitalized for longer than 90 days, were hospitalized in nursing hospitals, public health institutions, or clinics, were hospitalized when younger than 18 years old, and had the diagnosis code of ischemic stroke in the record within 1 year before hospitalization. Additionally, to focus on patients with noncardioembolic ischemic stroke who have indications for antiplatelet therapy, we excluded cases that had diagnosis records of atrial fibrillation, heart failure, myocardial infarction, or valvular heart disease for 1 year before hospitalization. Figure 1 illustrates our study population selection process.

**Figure 1.**
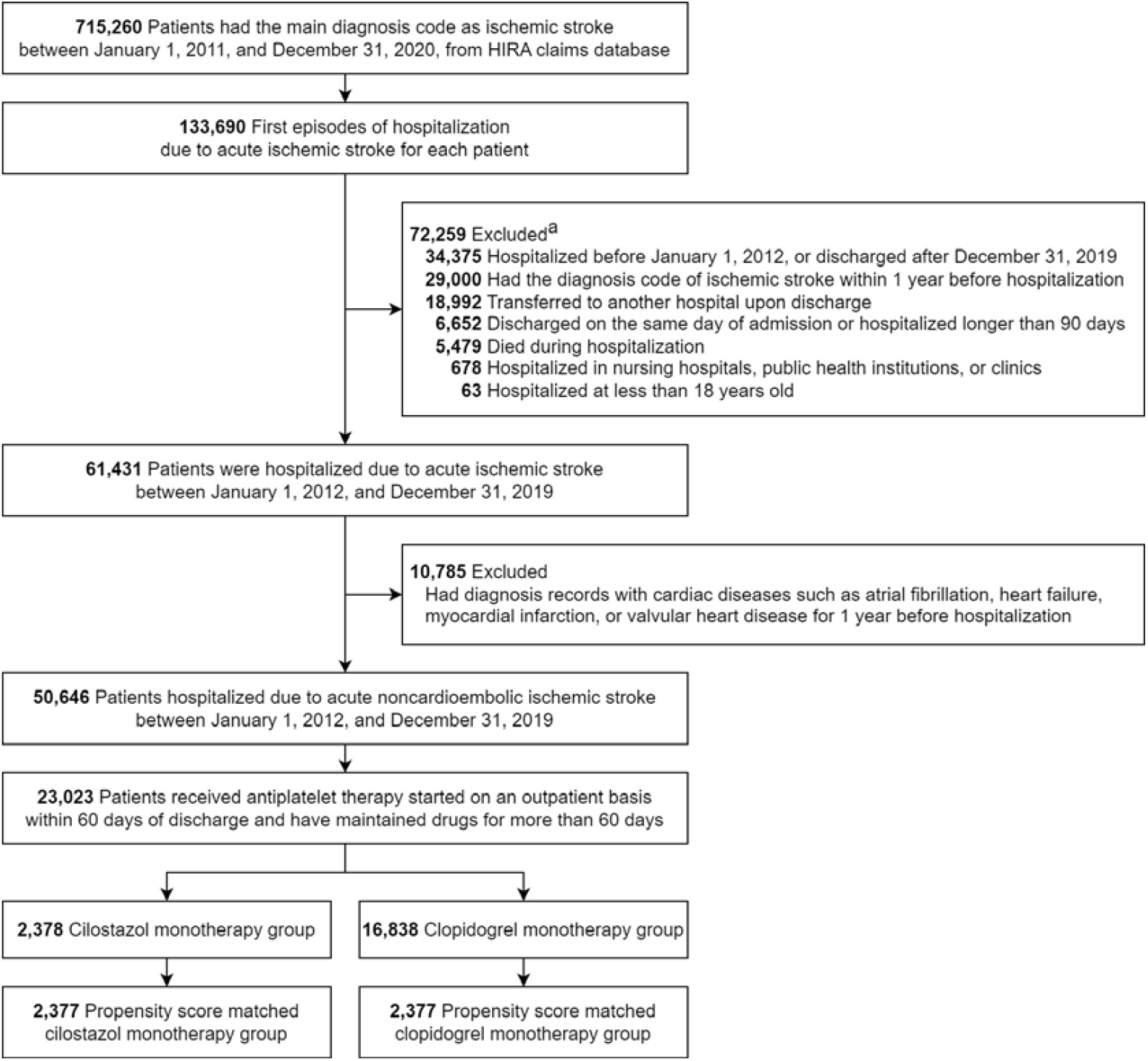
Study flowchart. a, Reasons for exclusion may include duplication.

### Study medications

From the selected patients, we observed those who received either cilostazol or clopidogrel monotherapy as an outpatient prescription within 60 days after discharge during the period of use of each drug. We considered continued administration if the grace period for taking the drug was less than 60 days. We terminated the observation when the prescription of the drug was discontinued, prescribed at intervals of more than 60 days, replaced by another antiplatelet agent, or added other antiplatelet agents during monotherapy. We excluded patients with a total antiplatelet treatment duration of fewer than 60 days, those who received oral anticoagulants at the time of hospitalization, or those who received study drugs in combination with oral anticoagulants or other antiplatelet agents.

### Outcomes

We defined the index date as the date of discharge from acute stroke hospitalization. The primary outcome was a recurrence of acute ischemic stroke, and the secondary outcomes were all-cause death, acute myocardial infarction, and hemorrhagic stroke. The composite outcome consisted of a recurrence of ischemic stroke, the occurrence of hemorrhagic stroke or myocardial infarction, and all-cause death. The safety outcome was major gastrointestinal (GI) bleeding. The operational definitions for each outcome are provided in Table S1 in the Supplement. We performed a time-to-event analysis, censoring when outcome events occurred, medical records were no longer available, the patient was recorded as deceased, study drugs were discontinued, other antiplatelet agents were added during monotherapy, or December 31, 2020, was reached.

### Subgroup analysis

As a subgroup analysis, we categorized the exposed groups by sex, the age group for those under 70 and 70 or more, hypertension, diabetes, vascular disease, dyslipidemia, previous TIA, previous ischemic hemorrhage, and previous GI bleeding and calculated incidence rates and HR with 95% CI in recurrent ischemic stroke, composite outcome and major GI bleeding.

### Statistical analysis

We utilized 1:1 propensity score matching (PSM) to balance the distribution of baseline characteristics between the two study groups. PSM applied logistic regression analysis using variables such as age, sex, length of hospital stays, type of medical institution, with or without EVT or IVT during hospitalization, hypertension, diabetes mellitus, dyslipidemia, vascular disease, previous cerebral hemorrhage, previous transient ischemic attack, and previous GI bleeding. We adopted the nearest neighbor matching algorithm and set the caliper as 0.2 times the standard deviation of the logit of the propensity score.

The baseline characteristics were summarized as median and interquartile ranges (IQRs) for continuous variables or frequency (percentage) for categorical variables. Categorical variables were compared using Chi-square or Fisher’s exact tests. Continuous variables with normal distributions were compared using the t-test, and those without normal distributions were compared using the Mann–Whitney U test.

After propensity score matching, the cumulative incidence was calculated as a percentage for each outcome, and the incidence rate was calculated as 1000PY using patient year (PY). Kaplan–Meier curves were generated for both the overall and matched cohorts. We compared the cumulative incidences between the two groups using the log-rank test. The hazard ratio with 95% CI between the two groups after propensity score matching was compared using a Cox proportional hazard model in which the study group was the independent variable and the matching pairs were stratified. The proportional hazards assumption was examined using the Schoenfeld residuals test and no significant violations were found in any of the outcomes.

All statistical analyses were conducted using the R software version 3.5.1 (R Foundation for Statistical Computing, Vienna, Austria; www.r-project.org). The “survival” package was used for survival analysis and the “MatchIt” package was used for the propensity score matching analysis. The R codes used for data handling and analysis were attached in Supplemental Methods. Statistical significance was set at 2-sided P < 0.05.

## Results

### Study population

Among 715,260 ischemic stroke patients from January 1, 2011, to December 31, 2020, extracted from the HIRA database, 133,690 patients were hospitalized for acute ischemic stroke. Finally, after applying the exclusion criteria, 50,646 patients were identified as those who were hospitalized and discharged due to acute noncardioembolic ischemic stroke between January 1, 2012, and December 31, 2019. Of these, 23,023 patients who began antiplatelet therapy within 60 days of discharge and continued for more than 60 days were selected as the target population (Figure 1).

The most commonly prescribed antiplatelet therapy was clopidogrel monotherapy for 16,838 patients (73.1%), followed by cilostazol monotherapy for 2,378 patients (10.3%) and aspirin/clopidogrel combination therapy for 1,968 patients (8.5%). Other antiplatelet therapies included monotherapy for 1,266 patients (5.5%), dual therapy for 533 patients (2.3%), and triple therapy for 40 patients (0.2%).

### Baseline characteristics

Before propensity score matching, the cilostazol group consisted of 2,378 patients and the clopidogrel group of 16,838 patients. Demographic and clinical characteristics of the patients at baseline are shown in Table 1. Both groups had a median (IQR) age of 67 (57-76) years with no significant difference between them (P = 0.36). The proportion of males was also similar, at 1,491 (62.7%) in the cilostazol group and 10,634 (63.2%) in the clopidogrel group (P = 0.68). However, there were significant differences in insurance type, type of medical institutions, and regional characteristics between the two groups. The cilostazol group had a higher frequency of hypertension, diabetes, dyslipidemia, or previous cerebral hemorrhage compared to the clopidogrel group (P < 0.05). Additionally, the proportion of patients who received EVT was higher in the clopidogrel group and the median length of hospital stay tended to be longer in the cilostazol group, while the median duration of drug use was longer in the clopidogrel group.

**Table 1.** Patient baseline characteristics before and after propensity score matching.

| Characteristics, n (%) | Before matching |  |  | After matching |  |  |
| --- | --- | --- | --- | --- | --- | --- |
|  | Cilostazol<br>(n=2,378) | Clopidogrel<br>(n=16,838) | P value | Cilostazol<br>(n=2,377) | Clopidogrel<br>(n=2,377) | P value |
| Sex |  |  | 0.68 |  |  | 0.23 |
| Male | 1491 (62.7) | 10634 (63.2) |  | 1491 (62.7) | 1450 (61.0) |  |
| Female | 887 (37.3) | 6204 (36.8) |  | 886 (37.3) | 927 (39.0) |  |
| Age (years, median [1QR;<br>3QR]) | 67.0 [57.0; 76.0] | 67.0 [57.0; 76.0] | 0.36 | 67.0 [57.0; 76.0] | 67 [58.0; 76.0] | 0.74 |
| Age group |  |  | 0.49 |  |  | 0.93 |
| <40 | 55 (2.3) | 420 (2.5) |  | 55 (2.3) | 57 (2.4) |  |
| 40-69 | 1254 (52.7) | 9055 (53.8) |  | 1253 (52.7) | 1263 (53.1) |  |
| ≥70 | 1069 (45.0) | 7363 (43.7) |  | 1069 (45.0) | 1057 (44.5) |  |
| Insurance type |  |  | 0.001 |  |  | 0.03 |
| NHI | 2172 (91.3) | 15687 (93.2) |  | 2171 (91.3) | 2212 (93.1) |  |
| MedAid/PVI | 206 (8.7) | 1151 (6.8) |  | 206 (8.7) | 165 (6.9) |  |
| Type of medical<br>institution |  |  | <0.001 |  |  | 0.99 |
| Tertiary hospital | 648 (26.8) | 5636 (33.5) |  | 647 (26.8) | 638 (26.8) |  |
| General hospital | 1624 (68.3) | 10080 (59.9) |  | 1624 (68.3) | 1625 (68.4) |  |
| Hospital | 116 (4.9) | 1122 (6.7) |  | 116 (4.9) | 114 (4.8) |  |
| Region |  |  | <0.001 |  |  | <0.001 |
| Capital | 554 (23.3) | 3038 (18.0) |  | 554 (23.3) | 119 (18.9) |  |
| Metropolitan | 747 (31.4) | 5783 (34.3) |  | 747 (31.4) | 726 (30.5) |  |
| Others | 1077 (45.3) | 8017 (47.6) |  | 1077 (45.3) | 1202 (50.6) |  |
| Length of hospital stay<br>(days, median [1QR;<br>3QR]) | 9.0 [7.0; 15.0] | 9.0 [6.0; 14.0] | <0.001 | 9.0 [7.0; 15.0] | 9.0 [7.0; 15.0] | 0.64 |
| CT |  |  | 0.01 |  |  | 0.85 |
| No | 1572 (66.1) | 11563 (68.7) |  | 1572 (66.1) | 1565 (65.8) |  |
| Yes | 806 (33.9) | 5275 (31.3) |  | 805 (33.9) | 812 (34.2) |  |
| MRI |  |  | 0.08 |  |  | 0.73 |
| No | 1058 (44.5) | 7814 (46.4) |  | 1057 (44.5) | 1044 (43.9) |  |
| Yes | 1320 (55.5) | 9024 (53.6) |  | 1320 (55.5) | 1333 (56.1) |  |
| EVT |  |  | 0.005 |  |  | 0.78 |
| No | 2271 (95.5) | 15837 (94.1) |  | 2270 (95.5) | 2265 (95.3) |  |
| Yes | 107 (4.5) | 1001 (5.9) |  | 107 (4.5) | 112 (4.7) |  |
| IVT |  |  | 0.16 |  |  | 0.68 |
| No | 1646 (69.2) | 11410 (67.8) |  | 1645 (69.2) | 1659 (69.8) |  |
| Yes | 732 (30.8) | 5428 (32.2) |  | 732 (30.8) | 718 (30.2) |  |
| Hypertension |  |  | 0.005 |  |  | 0.16 |
| No | 1112 (46.8) | 8394 (49.9) |  | 1112 (46.8) | 1063 (44.7) |  |
| Yes | 1266 (53.2) | 8444 (50.1) |  | 1265 (53.2) | 1314 (55.3) |  |
| Diabetes mellitus |  |  | <0.001 |  |  | 0.41 |
| No | 1562 (65.7) | 12017 (71.4) |  | 1561 (65.7) | 1533 (64.5) |  |
| Yes | 816 (34.3) | 4821 (28.6) |  | 816 (34.3) | 844 (35.5) |  |
| Vascular disease |  |  | 0.06 |  |  | 0.91 |
| No | 1952 (82.1) | 14081 (83.6) |  | 1951 (82.1) | 1947 (81.9) |  |
| Yes | 426 (17.9) | 2757 (16.4) |  | 426 (17.9) | 430 (18.1) |  |
| Dyslipidemia |  |  | 0.007 |  |  | 0.61 |
| No | 1498 (63.0) | 11083 (65.8) |  | 1498 (63.0) | 1480 (62.3) |  |
| Yes | 880 (37.0) | 5755 (34.2) |  | 879 (37.0) | 897 (37.7) |  |
| Prior TIA |  |  | 0.2 |  |  | 0.94 |
| No | 2282 (96.0) | 16250 (96.5) |  | 2282 (96.0) | 2284 (96.1) |  |
| Yes | 96 (4.0) | 588 (3.5) |  | 95 (4.0) | 93 (3.9) |  |
| Hemorrhagic stroke |  |  | <0.001 |  |  | 0.68 |
| No | 2326 (97.8) | 16706 (99.2) |  | 2326 (97.8) | 2331 (98.1) |  |
| Yes | 52 (2.2) | 132 (0.8) |  | 51 (2.1) | 46 (1.9) |  |
| GI hemorrhage |  |  | 0.31 |  |  | 0.93 |
| No | 2321 (97.6) | 16369 (97.2) |  | 2320 (97.6) | 2331 (98.1) |  |
| Yes | 57 (2.4) | 469 (2.8) |  | 57 (2.4) | 59 (2.5) |  |
| Medication day (days,<br>median [1QR; 3QR]) | 605.5 [267.0;<br>1194.0] | 632.0 [309.0;<br>1273.0] | 0.004 | 605.5 [267.0;<br>1193.0] | 613.0 [304.0;<br>1246.0] | 0.15 |

Abbreviations: MedAid, Medical aid; PVI, Patriots & veterans' insurance; NHI, National health insurance; CT, computed tomography;
MRI, magnetic resonance imaging; IVT, intravenous thrombolytic therapy; EVT, endovascular treatment; TIA, transient ischemic attack;
GI, gastrointestinal

After propensity score matching, 2,377 patients were matched in each group. The baseline characteristics used as matching variables were comparable between the two study groups (P > 0.05). The standardized differences for the baseline variables were also less than 0.1 (Table S2 in Supplement).

### Clinical outcomes

The follow-up period for all outcomes ranged from a minimum of 90 days to a maximum of 8 years. The results of the clinical outcomes before and after propensity score matching were not statistically different between the two study groups (Table 2 and Figure 2). The Kaplan-Meier curves for the pre-matching population are presented in Figure S1 in Supplement. The cilostazol group had comparable effectiveness in preventing the recurrence of acute ischemic stroke compared with the clopidogrel group (cilostazol versus clopidogrel: 64 (2.7%) versus 75 (3.2%); HR, 0.87; 95% CI, 0.62-1.21; Figure 2A). Similarly, the incidence of all-cause death (1.9% vs 1.7%; HR, 1.19; 95% CI, 0.78-1.82; Figure 2B), acute myocardial infarction (0.4% vs 0.7%; HR, 0.60; 95% CI, 0.27-1.30; Figure 2C), and hemorrhagic stroke (0.3% vs 0.3%; HR, 1.02; 95% CI, 0.36-2.92; Figure 2D) were similar. The composite outcome, which synthesized these outcomes, showed comparable results (5.1% vs 5.5%; HR, 0.95; 95% CI, 0.75-1.22; Figure 2E). The safety outcome was comparable between the cilostazol group and the clopidogrel group (1.3% vs 1.5%; HR, 0.92; 95% CI, 0.57-1.47; Fig 2F).

**Table 2.** Incidence rates and hazard ratios for clinical outcomes according to cilostazol versus clopidogrel.

| Clinical outcomes | Incidence rates |  |  |  |  |  | Cox proportional hazards analysis |  |
| --- | --- | --- | --- | --- | --- | --- | --- | --- |
|  | Cilostazol (n = 2377) |  |  | Clopidogrel (n = 2377) |  |  | HR (95% CI) | P value |
|  | Event (%) | Years | 1000PY | Event (%) | Years | 1000PY |  |  |
| Recurrent AIS | 64 (2.7) | 5306 | 12.1 | 75 (3.2) | 5537 | 13.5 | 0.87 (0.62-1.21) | 0.41 |
| All-cause death | 46 (1.9) | 5409 | 8.50 | 40 (1.7) | 5620 | 7.12 | 1.19 (0.78-1.82) | 0.43 |
| Myocardial infarction | 10 (0.4) | 5403 | 1.85 | 17 (0.7) | 5613 | 3.03 | 0.60 (0.27-1.30) | 0.2 |
| Hemorrhagic stroke | 7 (0.3) | 5408 | 1.29 | 7 (0.3) | 5620 | 1.25 | 1.02 (0.36-2.92) | 0.97 |
| Death, MI, or stroke | 121 (5.1) | 5299 | 2.28 | 130 (5.5) | 5531 | 2.35 | 0.95 (0.75-1.22) | 0.71 |
| Major GI bleeding | 31 (1.3) | 5359 | 5.78 | 35 (1.5) | 5591 | 6.26 | 0.92 (0.57-1.47) | 0.71 |

Hazard ratios are for the cilostazol group compared with the clopidogrel group in the matched population.
Abbreviations: CI, confidence interval; HR, hazard ratio; AIS, acute ischemic stroke; MI, myocardial infarction; GI, gastrointestinal

**Figure 2.**
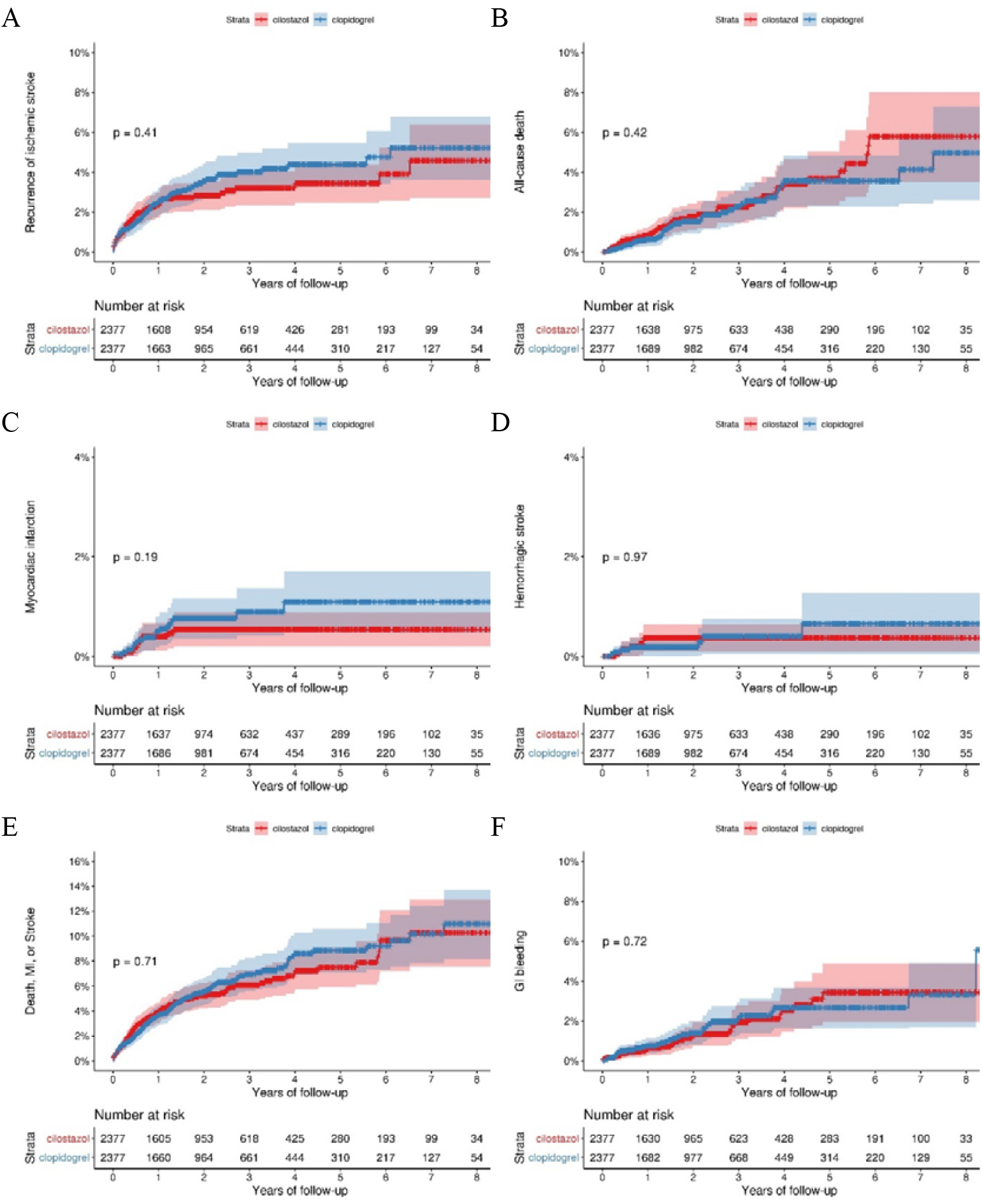
Kaplan-Meier curves for clinical outcomes in the matched population. The Kaplan-Meier curves for time to the first event of the primary outcome, defined as recurrent ischemic stroke (A), the secondary outcome of all-cause death (B), acute myocardial infarction (C), hemorrhagic stroke (D), and composite outcome consisting of occurrence for stroke, acute myocardial infarction, or death (E), and to the safety outcome of major gastrointestinal bleeding (F) are shown. P indicates the log-rank P value.

### Subgroup analysis

In the subgroup analysis, cilostazol appeared to be more effective than clopidogrel in preventing ischemic stroke recurrence in patients with hypertension (HR, 0.65; 95% CI, 0.41-1.01). The hazard ratio of composite outcome for cilostazol was 0.73 (95% CI, 0.52-1.01) in the hypertensive group. However, there were no statistically significant differences between the two drugs in the other subgroups (Figure S2 and Figure S3 in Supplement). Additionally, outcomes for major GI bleeding were comparable between the two drugs in all subgroups (Figure S4 in Supplement).

## Discussion

Our analysis of nationwide insurance claims data for up to 8 years in Korea found that cilostazol and clopidogrel were similarly effective and safe in patients with noncardioembolic ischemic stroke. In the primary outcome of recurrent ischemic stroke, the incidence tended to be lower in the cilostazol group compared to the clopidogrel group after 1 year from discharge, however, the difference was not statistically significant. While there was a trend towards a higher incidence of all-cause mortality in the cilostazol group after 5 years, there was no significant difference throughout the observation period. Both antiplatelet agents showed comparable incidences of major GI bleeding.

While not statistically significant, the tendency for cilostazol to show a lower incidence of recurrent ischemic stroke than clopidogrel 1 year after discharge may be related to the benefit of long-term use of cilostazol. A meta-analysis of the efficacy and safety of cilostazol in acute and chronic phases reported that cilostazol may be effective in the secondary prevention of the chronic phase of ischemic stroke.^18^ Additionally, another meta-analysis reported apparent benefits in reducing the recurrence of ischemic stroke when cilostazol was used for more than 6 months.^14^ However, we did not find a benefit of cilostazol in long-term use for all-cause mortality. This may have offset the effect on recurrence rates in the composite outcome. Therefore, it is difficult to prove the superiority of cilostazol compared to clopidogrel based on our results.

In a study comparing cilostazol and clopidogrel in patients with chronic ischemic stroke using Taiwanese insurance claims data for up to 3 years, Lee et al found that the effectiveness and safety of the two drugs were not significantly different.^19^ The study reported a hazard ratio of 0.89 (95% CI, 0.62-1.29) for ischemic stroke recurrence, 1.10 (95% CI, 0.78-1.55) in all-cause mortality, 0.97 (95% CI, 0.32-3.00) for myocardial infarction, 0.97 (95% CI, 0.38-2.48) for cerebral hemorrhage, and 0.85 (95% CI, 0.60-1.21) for gastrointestinal bleeding at a 3-year follow-up.^19^ These results were similar to our study in all outcomes except cerebral hemorrhage. A network meta-analysis study also showed cilostazol has a lower bleeding risk than clopidogrel, although it was not statistically significant.^20^ Despite suggestions that the pleiotropic effect of cilostazol may be effective in preventing hemorrhagic stroke based on its pharmacological mechanism, ^21^ it is not yet clear that cilostazol is different from clopidogrel. Based on the results of our study and previous studies, cilostazol may be a reasonable option as an antiplatelet agent for the secondary prevention of stroke, showing comparable effectiveness and safety to clopidogrel, at least in Asians.

Subgroup analysis revealed that cilostazol was more effective in preventing stroke recurrence in patients with hypertension, a finding supported by similar patterns in the study by Lee et al.^19^ The blood pressure-lowering effect of cilostazol may be attributed to its ability to promote endothelial vasodilation^22^ or decrease angiotensin II-induced apoptosis.^23^ In a clinical trial, the combination therapy of cilostazol with either aspirin or clopidogrel resulted in a 2 - 4 mmHg decrease in systolic blood pressure compared to monotherapy (P = 0.29).^24^ Subgroup analysis of the CSPS study also found that cilostazol was more effective in preventing stroke in patients with diabetes and/or hypertension compared to those without these conditions.^25^ However, there is limited data comparing cilostazol to clopidogrel in hypertensive patients, therefore, additional studies are needed to determine whether cilostazol is more beneficial than clopidogrel in hypertensive patients and the extent of its benefit.

Cilostazol is contraindicated in patients with heart failure due to its ability to inhibit PDE III, which can lead to positive inotropic effects.^26^ Long-term use of PDE III inhibitors has been linked to increased mortality from heart failure.^27^ A real-world study in Taiwan also found that cilostazol may be associated with an increased risk of heart failure in diabetic patients.^28^ As cilostazol is not approved for cardiogenic stroke, patients with heart disease such as heart failure were excluded from our investigation.

Clopidogrel is a prodrug that is converted into an active metabolite by cytochrome P450 (CYP) 2C19.^29^ Poor metabolizers of CYP2C19 are more common in Asians,^30^ and clopidogrel may not show sufficient efficacy in these individuals.^31^ A meta-analysis has reported that carriers of CYP2C19 genetic polymorphisms treated with clopidogrel are at increased risk of stroke and composite vascular events.^32^ However, the routine use of CYP2C19 genotyping before clopidogrel use is controversial.^33^ Alternative antiplatelet agents such as cilostazol, ticagrelor, and ticlopidine may be recommended for individuals who do not respond to clopidogrel.^33^ However, these alternative therapies also have their own set of contraindications and precautions. Therefore, clopidogrel and cilostazol should be chosen and modulated considering drug-drug interactions, and the patient’s genotype.

This study has several limitations. Firstly, the accuracy of diagnosis coding may be compromised by the nature of insurance claims data. Additionally, the severity of the patient’s disease could not be accurately determined as medical records were not accessible. Furthermore, the study was unable to differentiate between the various types of strokes, despite recent guidelines recommending different antiplatelet agents based on the type and severity of stroke.^3^ Additionally, there may be unmeasured confounding variables despite performing propensity score matching. Secondly, the absolute incidence rates of most outcomes in our study were somewhat lower than in previous studies.^19,34^ The lower incidence rate is likely due to differences in the operational definitions of outcome variables in this study. We only observed the subjects during the period of use of each antiplatelet agent, included the first event in each patient, and did not include events that occurred during hospitalization. Additionally, we excluded patients readmitted the next day after discharge or hospitalized for more than 90 days. Furthermore, our analysis of mortality outcomes was limited to deaths that occurred in medical institutions, as the insurance claims data used in this study only record deaths that occur in these settings. Thirdly, the study does not have information on the appropriateness of using monotherapy as the first regimen and the reasons for choosing cilostazol or clopidogrel. Lastly, the results of this study are limited to the Asian population and may not be generalize to other populations.

Despite these limitations, this study is meaningful as it is a nationwide, real-world study that examines the long-term effectiveness and safety of cilostazol and clopidogrel in noncardioembolic ischemic stroke patients. The study found that cilostazol, commonly used in Asian countries, is as effective and safe for long-term use in secondary stroke prevention as clopidogrel. Further research is needed to determine which antiplatelet agent is more advantageous based on the patient’s baseline characteristics.

## Data Availability

The data used for this study were obtained from the HIRA research database (M20220628003) and were stored on a separate server managed by the HIRA. All authors had full access to all the data in the study and take responsibility for its integrity and the data analysis.

## Non-standard Abbreviations and Acronyms

CASISP: Cilostazol versus Aspirin for Secondary Ischaemic Stroke Prevention
CSPS: Cilostazol for Prevention of Secondary Stroke
CT: computed tomography
CYP: cytochrome P450
DAPT: dual antiplatelet therapy
EVT: endovascular treatment
GI: gastrointestinal
HIRA: Health Insurance and Review Assessment
IVT: intravenous thrombolytic therapy
MRI: magnetic resonance imaging
PDE: phosphodiesterase
PICASSO: Prevention of Cardiovascular Events in Ischemic Stroke Patients with High Risk of Cerebral Hemorrhage
PSM: propensity score matching
SAPT: single antiplatelet therapy
TIA: transient ischemic attack

## Authors’ contributions

YJL and NKJ had full access to all of the data in the study and take responsibility for the integrity of the data and the accuracy of the data analysis. YJL and NKJ conceived and designed the study; YJL performed the analysis; YJL first drafted the manuscript; YJL and NKJ participated in drafting the article and approved the final version to be submitted for publication.

## References

1. Sacco RL, Wolf PA, Kannel WB, McNamara PM. Survival and recurrence following stroke. The Framingham study. Stroke. 1982;13:290–295. doi: 10.1161/01.str.13.3.290

2. Dawson J, Béjot Y, Christensen LM, De Marchis GM, Dichgans M, Hagberg G, Heldner MR, Milionis H, Li L, Pezzella FR, et al. European Stroke Organisation (ESO) guideline on pharmacological interventions for long-term secondary prevention after ischaemic stroke or transient ischaemic attack. European Stroke Journal. 2022;7:I–XLI. doi: 10.1177/23969873221100032

3. Kleindorfer DO, Towfighi A, Chaturvedi S, Cockroft KM, Gutierrez J, Lombardi-Hill D, Kamel H, Kernan WN, Kittner SJ, Leira EC, et al. 2021 Guideline for the Prevention of Stroke in Patients With Stroke and Transient Ischemic Attack: A Guideline From the American Heart Association/American Stroke Association. Stroke. 2021;52:e364–e467. doi: 10.1161/STR.0000000000000375

4. Clinical Practice Guideline for Stroke. In: Seoul, KR: The writing group of clinical practice guideline for stroke; 2016.

5. Committee CS. A randomised, blinded, trial of clopidogrel versus aspirin in patients at risk of ischaemic events (CAPRIE). CAPRIE Steering Committee. Lancet. 1996;348:1329–1339. doi: 10.1016/s0140-6736(96)09457-3

6. Paciaroni M, Ince B, Hu B, Jeng JS, Kutluk K, Liu L, Lou M, Parfenov V, Wong KSL, Zamani B, et al. Benefits and Risks of Clopidogrel vs. Aspirin Monotherapy after Recent Ischemic Stroke: A Systematic Review and Meta-Analysis. Cardiovasc Ther. 2019;2019:1607181. doi: 10.1155/2019/1607181

7. McQuaid KR, Laine L. Systematic review and meta-analysis of adverse events of low-dose aspirin and clopidogrel in randomized controlled trials. Am J Med. 2006;119:624–638. doi: 10.1016/j.amjmed.2005.10.039

8. Sorkin EM, Markham A. Cilostazol. Drugs Aging. 1999;14:63–71; discussion 72-63. doi: 10.2165/00002512-199914010-00005

9. de Havenon A, Sheth KN, Madsen TE, Johnston KC, Turan TN, Toyoda K, Elm JJ, Wardlaw JM, Johnston SC, Williams OA, et al. Cilostazol for Secondary Stroke Prevention: History, Evidence, Limitations, and Possibilities. Stroke. 2021;52:e635–e645. doi: 10.1161/strokeaha.121.035002

10. Huang Y, Cheng Y, Wu J, Li Y, Xu E, Hong Z, Li Z, Zhang W, Ding M, Gao X, et al. Cilostazol as an alternative to aspirin after ischaemic stroke: a randomised, double-blind, pilot study. Lancet Neurol. 2008;7:494–499. doi: 10.1016/s1474-4422(08)70094-2

11. Shinohara Y, Katayama Y, Uchiyama S, Yamaguchi T, Handa S, Matsuoka K, Ohashi Y, Tanahashi N, Yamamoto H, Genka C, et al. Cilostazol for prevention of secondary stroke (CSPS 2): an aspirin-controlled, double-blind, randomised non-inferiority trial. Lancet Neurol. 2010;9:959–968. doi: 10.1016/S1474-4422(10)70198-8

12. Kim BJ, Lee EJ, Kwon SU, Park JH, Kim YJ, Hong KS, Wong LKS, Yu S, Hwang YH, Lee JS, et al. Prevention of cardiovascular events in Asian patients with ischaemic stroke at high risk of cerebral haemorrhage (PICASSO): a multicentre, randomised controlled trial. Lancet Neurol. 2018;17:509–518. doi: 10.1016/s1474-4422(18)30128-5

13. Kamal AK, Naqvi I, Husain MR, Khealani BA. Cilostazol versus aspirin for secondary prevention of vascular events after stroke of arterial origin. Cochrane database of systematic reviews. 2011;2011:CD008076. doi: 10.1002/14651858.CD008076.pub2

14. McHutchison C, Blair GW, Appleton JP, Chappell FM, Doubal F, Bath PM, Wardlaw JM. Cilostazol for Secondary Prevention of Stroke and Cognitive Decline: Systematic Review and Meta-Analysis. Stroke. 2020;51:2374–2385. doi: 10.1161/strokeaha.120.029454

15. Tan CH, Wu AG, Sia C-H, Leow AS, Chan BP, Sharma VK, Yeo LL, Tan BY. Cilostazol for secondary stroke prevention: systematic review and meta-analysis. Stroke and Vascular Neurology. 2021;6:410–423. doi: 10.1136/svn-2020-000737

16. Kim J-A, Yoon S, Kim L-Y, Kim D-S. Towards Actualizing the Value Potential of Korea Health Insurance Review and Assessment (HIRA) Data as a Resource for Health Research: Strengths, Limitations, Applications, and Strategies for Optimal Use of HIRA Data. Journal of Korean Medical Science. 2017;32:718. doi: 10.3346/jkms.2017.32.5.718

17. Kim JY, Lee K-J, Kang J, Kim BJ, Han M-K, Kim S-E, Lee H, Park J-M, Kang K, Lee SJ, et al. Development of stroke identification algorithm for claims data using the multicenter stroke registry database. PLOS ONE. 2020;15:e0228997. doi: 10.1371/journal.pone.0228997

18. Shi L, Pu J, Xu L, Malaguit J, Zhang J, Chen S. The efficacy and safety of cilostazol for the secondary prevention of ischemic stroke in acute and chronic phases in Asian population--an updated meta-analysis. BMC Neurol. 2014;14:251. doi: 10.1186/s12883-014-0251-7

19. Lee T-H, Lin Y-S, Liou C-W, Lee J-D, Peng T-I, Liu C-H. Comparison of long-term efficacy and safety between cilostazol and clopidogrel in chronic ischemic stroke: a nationwide cohort study. Therapeutic advances in chronic disease. 2020;11:2040622320936418. doi: 10.1177/2040622320936418

20. Niu P-P, Guo Z-N, Jin H, Xing Y-Q, Yang Y. Antiplatelet regimens in the longterm secondary prevention of transient ischaemic attack and ischaemic stroke: an updated network meta-analysis. BMJ Open. 2016;6:e009013. doi: 10.1136/bmjopen-2015-009013

21. Takagi T, Hara H. Protective effects of cilostazol against hemorrhagic stroke: Current and future perspectives. J Pharmacol Sci. 2016;131:155–161. doi: 10.1016/j.jphs.2016.04.023

22. Nakamura K, Ikomi F, Ohhashi T. Cilostazol, an inhibitor of type 3 phosphodiesterase, produces endothelium-independent vasodilation in pressurized rabbit cerebral penetrating arterioles. J Vasc Res. 2006;43:86–94. doi: 10.1159/000089723

23. Shi MQ, Su FF, Xu X, Liu XT, Wang HT, Zhang W, Li X, Lian C, Zheng QS, Feng ZC. Cilostazol suppresses angiotensin II-induced apoptosis in endothelial cells. Mol Med Rep. 2016;13:2597–2605. doi: 10.3892/mmr.2016.4881

24. Toyoda K, Uchiyama S, Yamaguchi T, Easton JD, Kimura K, Hoshino H, Sakai N, Okada Y, Tanaka K, Origasa H, et al. Dual antiplatelet therapy using cilostazol for secondary prevention in patients with high-risk ischaemic stroke in Japan: a multicentre, open-label, randomised controlled trial. Lancet Neurol. 2019;18:539–548. doi: 10.1016/s1474-4422(19)30148-6

25. Shinohara Y, Gotoh F, Tohgi H, Hirai S, Terashi A, Fukuuchi Y, Otomo E, Itoh E, Matsuda T, Sawada T, et al. Antiplatelet cilostazol is beneficial in diabetic and/or hypertensive ischemic stroke patients. Subgroup analysis of the cilostazol stroke prevention study. Cerebrovasc Dis. 2008;26:63–70. doi: 10.1159/000135654

26. Kanlop N, Chattipakorn S, Chattipakorn N. Effects of cilostazol in the heart. Journal of Cardiovascular Medicine. 2011;12:88–95. doi: 10.2459/JCM.0b013e3283439746

27. Amsallem E, Kasparian C, Haddour G, Boissel JP, Nony P. Phosphodiesterase III inhibitors for heart failure. Cochrane Database Syst Rev. 2005;2005:Cd002230. doi: 10.1002/14651858.CD002230.pub2

28. Wu CK, Lin JW, Wu LC, Chang CH. Risk of Heart Failure Hospitalization Associated With Cilostazol in Diabetes: A Nationwide Case-Crossover Study. Front Pharmacol. 2018;9:1467. doi: 10.3389/fphar.2018.01467

29. Sangkuhl K, Klein TE, Altman RB. Clopidogrel pathway. Pharmacogenet Genomics. 2010;20:463–465. doi: 10.1097/FPC.0b013e3283385420

30. Scott SA, Sangkuhl K, Stein CM, Hulot JS, Mega JL, Roden DM, Klein TE, Sabatine MS, Johnson JA, Shuldiner AR. Clinical Pharmacogenetics Implementation Consortium guidelines for CYP2C19 genotype and clopidogrel therapy: 2013 update. Clin Pharmacol Ther. 2013;94:317–323. doi: 10.1038/clpt.2013.105

31. Lo C, Nguyen S, Yang C, Witt L, Wen A, Liao TV, Nguyen J, Lin B, Altman RB, Palaniappan L. Pharmacogenomics in Asian Subpopulations and Impacts on Commonly Prescribed Medications. Clin Transl Sci. 2020;13:861–870. doi: 10.1111/cts.12771

32. Pan Y, Chen W, Xu Y, Yi X, Han Y, Yang Q, Li X, Huang L, Johnston SC, Zhao X, et al. Genetic Polymorphisms and Clopidogrel Efficacy for Acute Ischemic Stroke or Transient Ischemic Attack: A Systematic Review and Meta-Analysis. Circulation. 2017;135:21–33. doi: 10.1161/circulationaha.116.024913

33. Lee CR, Luzum JA, Sangkuhl K, Gammal RS, Sabatine MS, Stein CM, Kisor DF, Limdi NA, Lee YM, Scott SA, et al. Clinical Pharmacogenetics Implementation Consortium Guideline for CYP2C19 Genotype and Clopidogrel Therapy: 2022 Update. Clin Pharmacol Ther. 2022;112:959–967. doi: 10.1002/cpt.2526

34. Chi NF, Wen CP, Liu CH, Li JY, Jeng JS, Chen CH, Lien LM, Lin CH, Sun Y, Chang WL, et al. Comparison Between Aspirin and Clopidogrel in Secondary Stroke Prevention Based on Real-World Data. J Am Heart Assoc. 2018;7:e009856. doi: 10.1161/jaha.118.009856

